# Direct RNA sequencing reveals an m^6^A downregulated on *APOEϵ4* RNA in brain from Alzheimer’s disease cases and controls

**DOI:** 10.64898/2026.11.21.26368646

**Authors:** Grant A. Fox, Madeline L. Page, Mark E. Wadsworth, Bernardo Aguzzoli Heberle, Lacey A. Gordon, David W. Fardo, Lance A. Johnson, Thomas G. Beach, Geidy E. Serrano, Peter T. Nelson, Mark T. W. Ebbert

## Abstract

*APOE* harbors the strongest common genetic risk factors for late-onset Alzheimer’s disease (AD), yet its disease mechanism remains incompletely understood. Two missense SNPs jointly define the *APOE* alleles, with rs429358 distinguishing ϵ4 from ϵ3, but whether other *APOE* features, such as mRNA modifications, contribute to *APOE*-associated AD risk is unknown. Here, we performed deep long-read direct RNA sequencing on human frontal cortex from six AD cases and six controls to characterize native mRNA modifications transcriptome-wide and across AD-relevant genes. We evaluated differential nucleotide modifications between cases and controls transcriptome-wide, and we then performed two association tests within *APOE*: (1) associations between mRNA modifications and rs429358 alleles in five ϵ3/ϵ4 heterozygotes; and (2) pairwise associations between co-occurring mRNA modifications. We identified four modifications associated with rs429358—all within 15 nucleotides of the SNP. Among them was an N6-methyladenosine (m^6^A) located at chr19:44,908,679 (OR = 0.53; p = 1.4 × 10^−109^)—five nucleotides upstream of rs429358—that was 14.5 percentage points lower on ϵ4-derived mRNA (27.7%) compared to ϵ3 (42.2%). This directional difference was observed within each individual, supporting a consistent allele-associated modification pattern (p = 4.2 × 10^−5^). We also identified 344 associations among 73 *APOE* modifications, suggesting a potentially complex, coordinated network of mRNA biological signals within *APOE*, alone. Our findings suggest that mRNA modifications may contribute to *APOE*-associated risk, though validation in large independent cohorts and functional experiments will be necessary to determine the biological consequences of these associations and their potential relevance to AD pathogenesis. Together, our findings highlight the potential of allele-resolved direct RNA sequencing to characterize genetic associations with RNA modifications at a major AD risk locus.

## Introduction

Apolipoprotein E (*APOE*) was the first gene associated with late-onset Alzheimer’s disease (AD) in 1993^1–3^ and remains the top known genetic risk factor with an odds ratio (OR) and 95% confidence interval (CI) of 3.0 (per ϵ4 allele) and 2.95-3.05 (p < 1 × 10^−300^)^4–6^, respectively. *APOE* has three primary haplotypes or alleles known as ϵ2, ϵ3, and ϵ4, where *APOE*ϵ4 is considered the risk allele while *APOE*ϵ2 and *APOE*ϵ3 have historically been considered protective and neutral, respectively— though Williams et al.^7^ make a strong argument that *APOE*ϵ3 should also be considered a risk allele. A fourth allele, known as either *APOE*ϵ1Y^8^ or *APOE*ϵ3r (ϵ3 in reverse)^9^, also exists but is rare with an allele frequency (AF) of 0.0001^10^ and is largely ignored. Few studies have estimated the population attributable fraction (PAF) for *APOE*ϵ4 in a true population-based AD cohort, but Williams et al. estimated that AD cases would diminish by 75.7% if both the *APOE*ϵ3 and *APOE*ϵ4 alleles could be replaced by *APOE*ϵ2 in the population, where approximately 31% and 44% were attributed to ϵ3 and ϵ4, respectively^11^. With an estimated 32 million people currently suffering from AD worldwide^12^, that would equate to protecting between 9.9 (replacing ϵ3), 14.1 (replacing ϵ4), or 24.2 (replacing both) million people (and their loved ones) from the effects of this devastating disease, at present.

Given *APOE*’s clear impact on AD risk, researchers have been studying its disease mechanism for decades, resulting in approximately 16,917 publications related to *APOE* and AD since its discovery. Despite these many studies, *APOE*’s role in disease development and progression remain incompletely understood. There are many hypotheses surrounding how *APOE* contributes to disease development, including amyloid-beta (Aβ) aggregation and clearance^13–19^, tau pathology^17,20–23^, lipid cholesterol and metabolism^19,24–30^, neuroinflammation and microglial dysfunction^21,26–36^, and blood-brain barrier dysfunction^37–39^, among others^40–43^. A recent genetic study by Shade et al.^6^ directly supports the Aβ and tau hypotheses, demonstrating that *APOE* is directly genetically associated with three AD-related neuropathologies (Aβ plaques including diffuse plaques, CERAD score for neuritic amyloid plaques, & Braak staging^44,45^), among other non-AD-specific associations. It is possible *APOE* would also associate with the other major hypotheses described in human neuropathology studies, but there are currently no clear neuropathological measurements available at autopsy to test association. Regardless, as described, numerous experimental studies demonstrate that *APOE* plays a major role in disease. Thus, fully understanding *APOE*’s AD mechanisms is essential to understanding disease etiology, development, and progression.

*APOE* is one of the few AD genes with genetic mutations that have clear functional conse-quences, providing clear mechanisms through which researchers can directly study its involvement in disease. Specifically, two non-synonymous SNPs—rs429358 (chr19:44,908,684; T→C; Cys112Arg) and rs7412 (chr19:44,908,822; C→T; Arg158Cys)—lie 138 nucleotides apart in exon 4 (**Fig. 1a**) and jointly define four possible APOE haplotypes: ϵ2 (T–T; Cys-Cys), ϵ3 (T–C; Cys-Arg), ϵ4 (C–C; Arg-Arg), and the rare ϵ3r (C–T; Arg-Cys; **Fig. 1b**). Relative to ϵ3, ϵ4 differs at rs429358 and ϵ2 differs at rs7412, producing amino acid substitutions whose biological consequences can be experimentally studied in myriad ways. The minor alleles rs429358-C (15%) and rs7412-T (8%) are relatively common in the UK Biobank and All of Us cohorts (**Fig. 1b**)^10,46,47^, but this can vary substantially by ancestry. The corresponding *APOE* haplotype frequencies and AD odds ratios (OR) vary (relative to ϵ3): ϵ2 (AF = 8%; OR = 0.70), ϵ3 (AF = 77%; OR = 1.0), ϵ4 (AF = 15%; OR = 3.62), and *APOE*ϵ3r (AF = 0.01%; OR = unknown). Given these variants are exonic and non-synonymous, they have direct implications on *APOE*’s downstream protein biology and could have additional unknown impacts on *APOE*’s mRNA biology.

**Figure 1.**
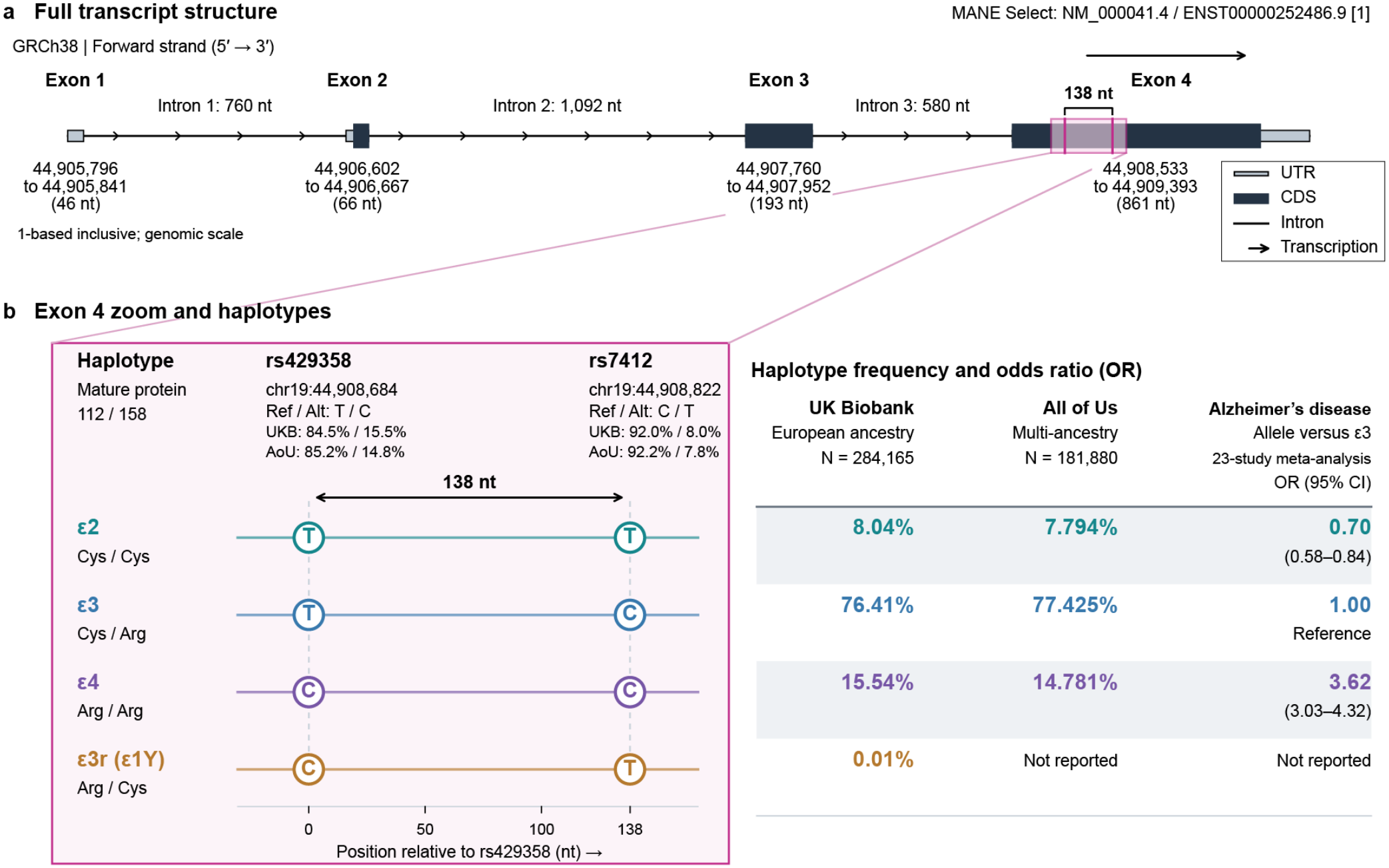
Schematic summarizing *APOE* SNP alleles and haplotypes. (**a**) The two *APOE* haplotype-defining SNPs, rs429358 and rs7412, are located 138 nucleotides apart in exon 4. (**b**) Jointly, rs429358 and rs7412 can result in four haplotypes known as ϵ2 (T-T), ϵ3 (T-C), ϵ4 (C-C), and the rare haplotype known as either ϵ1Y or ϵ3r (ϵ3 in reverse; C-T) because it is opposite the ϵ3 haplotype at both the DNA (ϵ3: T-C; ϵ3r: C-T) and protein (ϵ3: Cys-Arg; ϵ3r: Arg-Cys) levels. Thus, rs429358-C distinguishes ϵ4 from ϵ3, whereas rs7412-T distinguishes ϵ2 from ϵ3. The rare ϵ3r haplotype carries both substitutions. For reference, both the SNP and haplotype allele frequencies from the UK Biobank and All of Us are provided, as are the haplotype odds ratios (relative to the ϵ3 allele). Script used to create figure was generated by ChatGPT and human verified.

RNA modifications are key biological signals and an understudied area of mRNA biology, including in relation to *APOE* and its involvement in AD. Researchers have already discovered approximately 170 RNA modifications across all of biology^48–50^, with between 43^51^ and 50+^50^ reported in humans; of those, approximately ten types have been reported on human mRNA, including (1) N6-methyladenosine (m^6^A)^52,53^; (2) N1-methyladenosine (m^1^A)^54^; (3) 5-methylcytidine (m^5^C)^55^; (4) N4-acetylcytidine (ac^4^C)^56^; (5) ‘adenosine to inosine’ (A→I) editing^57,58^; (6) ‘Cytidine to uridine’ (C→U)^59,60^ editing; (7) pseudouridine (Ψ)^61,62^; (8) dihydrouridine (D)^63,64^; (9) 2’-O-methyl ribose on canonical nucleosides (Nm)^65^; and (10) N7-methylguanosine (m^7^G)^66,67^. Though its natural existence was originally in doubt^68–70^, m^7^G, which is also the canonical mRNA cap attached to the first transcribed nucleotide^71^, has only recently been established as a tenth internal mRNA modification.

Unfortunately, studying RNA modifications has been challenging for technological reasons because, in short, no technology has historically been able to directly measure RNA modifications with nucleotide-level, single-molecule resolution within their full RNA sequence context. Oxford Nanopore Technologies’ (ONT) direct RNA sequencing (dRNAseq) is a relatively new approach that holds the promise to simultaneously measure all RNA modifications—jointly with exonic genomic variants and RNA isoform structure, among other RNA biology features (e.g., polyadenylation). At present, however, ONT’s dRNAseq can only identify five basic modification types because machine learning models for the others do not yet exist. The five currently discoverable via dRNAseq include m^6^A, m^5^C, inosine, pseudouridine, and 2’-O-methyl ribose on canonical nucleosides (Nm). While dRNAseq is new and likely has important limitations, new technologies often reveal new biology, as shown by several dRNAseq studies in recent years^72–84^.

Here, as a proof-of-principle study, we performed deep dRNAseq in human prefrontal cortex (Brodmann Area 9/46) in a small cohort of AD cases (six) and controls (six). We first broadly characterized the land-scape of mRNA modifications in this cohort, both transcriptome-wide and across disease-relevant gene sets and then performed differential nucleotide modification analysis between cases and controls transcrip-tome-wide. We then performed two association tests within *APOE*: (1) associations between RNA modifications and exonic genetic variants (modification-variant associations) or RNA modification quantitative trait loci (rmQTL) within *APOE*; and (2) associations between co-occurring RNA modifications (modification-modification associations) within *APOE*. The latter two approaches are only possible because of the nucleotide-level, single-molecule resolution provided by ONT’s dRNAseq. Among other preliminary findings, we report a new, high-confidence association between the *APOE*ϵ4 allele and an m^6^A at position 44,908,679 on chromosome 19 (OR [95% CI] = 0.53 [0.50, 0.56]; p = 1.4 × 10^−109^), which is five nucleotides upstream of rs429358; the m^6^A is consistently downregulated on mRNA molecules originating from the *APOE*ϵ4 allele compared to molecules originating from *APOE*ϵ3 in five *APOE*ϵ3/ϵ4 heterozygotes. We are currently validating these results in a larger cohort.

## Results

We performed deep dRNAseq on six AD cases and six sex and age-matched controls (**Table 1**) using two PromethION flow cells (FLO-PRORNA004) per sample, resulting in an average of 22.93 ± 4.07 million aligned and usable reads per sample. With this sequencing depth, requiring a minimum counts-per-million (CPM) threshold of 1 for at least three samples, we observed expression for 31,848 RNA isoforms across 16,588 genes in at least three samples (**Fig. 2a**). In this study, we report sites for five modification types, including m^6^A, m^5^C, inosine, pseudouridine, and 2′-O-methyl ribose (Nm); Nm modifications can be present on adenosine (Am), cytidine (Cm), guanosine (Gm), and uridine (Um).

**Table 1.** Demographic, neuropathological, tissue-quality, and sequencing characteristics of the DLPFC cohort. Characteristics of the twelve dorsolateral prefrontal cortex tissue specimens (Brodmann area 9/46) retained for analysis are shown stratified by case or control. The cohort included six female and six male samples, with three of each sex per group. Continuous variables are reported as mean ± standard deviation (SD); categorical variables are reported as n. Sequencing metrics are summarized across per-sample values; yield is reported in gigabases (Gb) and read length in nucleotides (nt). No sequencing metric differed significantly between cases and controls (Welch’s t-test and Mann–Whitney U test; Benjamini–Hochberg-adjusted q < 0.05 for all metrics). *Differed significantly between cases and controls after Benjamini–Hochberg correction (Braak stage, p = 0.002, q = 0.017) ^†^Nominally significant but not after correction (p = 0.048, q = 0.210).

| **Characteristic** | **Cases** | **Controls** |
|---|---|---|
| **Subjects (n)** | 6 | 6 |
| **Sex (n)** |  |  |
| Male | 3 | 3 |
| Female | 3 | 3 |
| **Hispanic (n)** |  |  |
| Hispanic | 4 | 4 |
| Unknown | 2 | 2 |
| **Age at death (years; mean ± SD)** | 78.2 ± 6.9 | 79.7 ± 7.9 |
| **Postmortem interval (hours; mean ± SD)** | 2.8 ± 0.5 | 3.1 ± 0.8 |
| **<sup>†</sup>Brain weight (grams; mean ± SD)** | 1071.5 ± 138.6 | 1220.2 ± 64.7 |
| ***APOE*** **diplotype (n)** |  |  |
| ϵ3/ϵ3 | 3 | 4 |
| ϵ3/ϵ4 | 3 | 2 |
| **\*Braak stage (n)** |  |  |
| I | — | 3 |
| II | — | 1 |
| III | — | 2 |
| V | 6 | — |
| **Average RNA Quality Number (RQN; mean ± SD)** | 9.0 ± 0.6 | 9.2 ± 0.3 |
| **Average aligned reads (millions; mean ± SD)** | 24.39 ± 4.75 | 21.48 ± 2.98 |
| **Total yield (Gb; mean ± SD)** | 15.72 ± 5.22 | 13.20 ± 2.82 |
| **Aligned yield (Gb; mean ± SD)** | 14.70 ± 5.05 | 12.32 ± 2.75 |
| **Bases aligned (%; mean ± SD)** | 93.18 ± 1.45 | 93.26 ± 1.10 |
| **Median read length (nt; mean ± SD)** | 477.83 ± 115.81 | 459.83 ± 77.50 |
| **Read length N50 (nt; mean ± SD)** | 865.17 ± 205.85 | 841.33 ± 137.52 |

**Figure 2.**
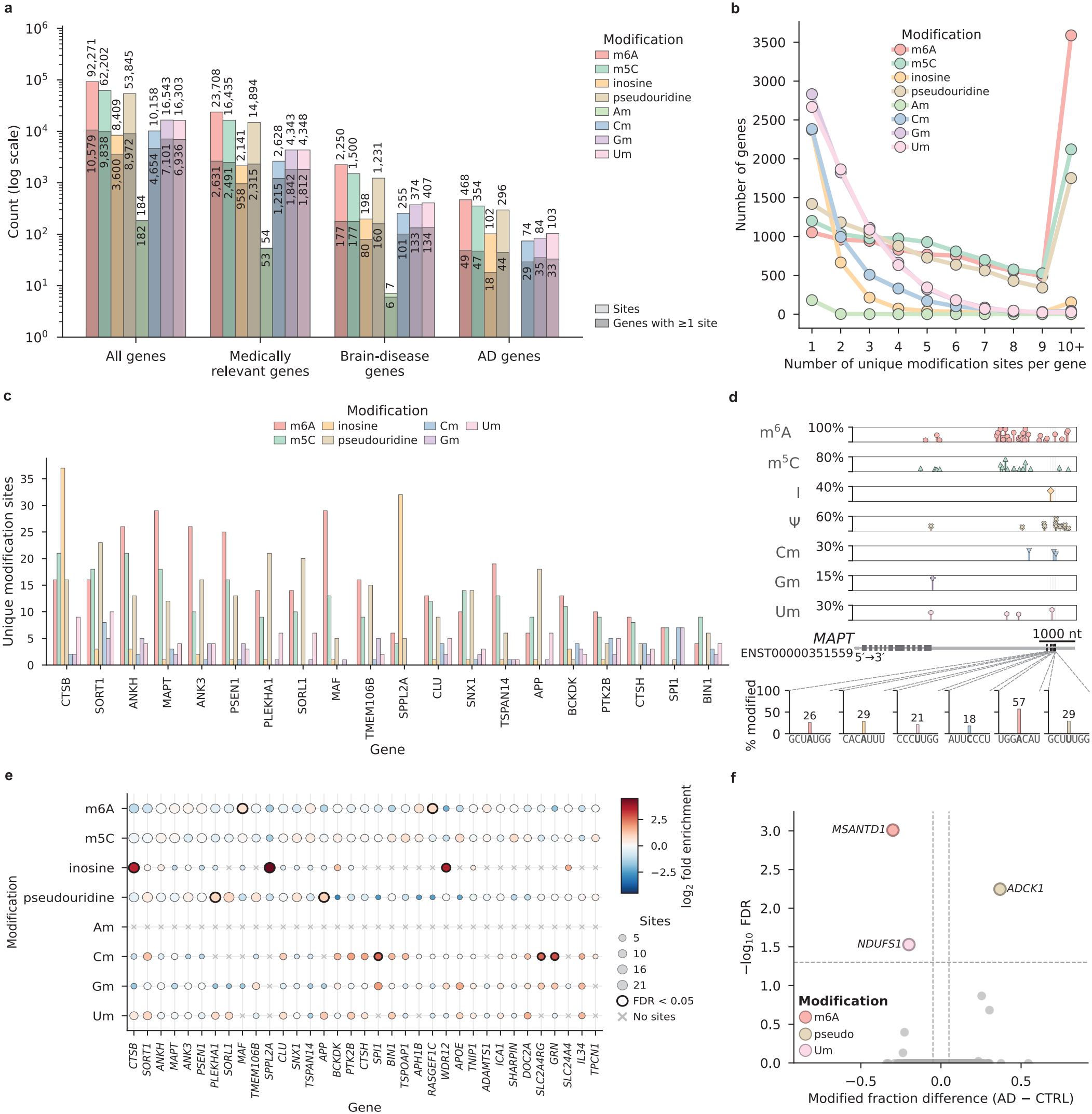
Transcriptome-wide landscape of mRNA modifications in human prefrontal cortex across Alzheimer’s disease genes. Direct RNA sequencing of six AD cases and six controls; modification sites required ≥30 reads of coverage and ≥10% modification rate in ≥6 of 12 samples. (**a**) Number of sites and genes with ≥1 site for each modification type across all, medically relevant, brain-disease, and AD-associated genes (log scale). (**b**) Number of genes by unique sites per gene (1 to 10+) for each modification type. (**c**) Unique modification sites in each of the 51 modified AD-associated genes, ranked by total sites and colored by modification type. (**d**) Percent of reads modified at sites along *MAPT* (ENST00000351559) for six closely clustered sites within 250 nucleotides in the 3` UTR with percent modified and ±3 nucleotides around the centered on modified site. Thick boxes, CDS; thin boxes, UTRs; introns are thin black lines; m^6^A, N6-methyladenosine; m^5^C, 5-methylcytidine; I, inosine; Ψ, pseudouridine; Am, Cm, Gm, and Um, 2′-O-methyladenosine, -cytidine, -guanosine, and -uridine. (**e**) Enrichment of each modification type within each AD-associated gene relative to transcriptome-wide (log_2_ fold enrichment); dot size indicates number of sites, with significant enrichment outlined (one-sided Fisher’s exact test, Benjamini–Hochberg FDR < 0.05). (**f**) Volcano plot showing three mRNA modifications associated with disease status, including one m^6^A in *MSANTD1*, one pseudouridine in *ADCK1*, and one Um in *NDUFS1*. Scripts used to create figures were generated by ChatGPT and human verified.

### Transcriptome-wide characterization of mRNA modifications

We identified 347,850 total mRNA modification sites transcriptome-wide that met our inclusion criteria of ≥30 read coverage with ≥10% modification rate in at least six of the twelve samples. Of these, 4,474 sites were in RNA arising from genomic locations lacking gene annotations (i.e., intergenic) and 83,461 sites overlapped more than one annotated gene; the remaining 259,915 sites mapped to a single gene, spanning 11,198 unique genes (76,987 unique RNA isoforms). Of the 259,915 modifications that mapped to a single gene, 92,271 (35.5%) were m^6^A (10,579 genes; 64,786 iso-forms), 62,202 (23.9%) were m^5^C (9,838 genes; 62,919 isoforms), 53,845 (20.7%) were pseudouridine (8,972 genes; 55,502 isoforms), 8,409 (3.2%) were inosine (3,600 genes; 15,761 isoforms), and 43,188 (16.6%) were Nm, comprised of 184 Am, 10,158 Cm, 16,543 Gm, and 16,303 Um sites (**Fig. 2a**). Am modifications appear to be rarer than the others.

We next examined these sites across three disease-relevant gene sets (**Fig. 2a**). In medically relevant genes^85,86^, 2,787 carried ≥1 modification site, for a total of 68,551 sites, including 23,708 m^6^A, 16,435 m^5^C, 14,894 pseudouridine, 2,141 inosine, and 11,373 Nm (54 Am, 2,628 Cm, 4,343 Gm, and 4,348 Um). For brain-disease associated genes^86^, 187 contained ≥1 site and accounted for 6,222 sites (2,250 m^6^A, 1,500 m^5^C, 1,231 pseudouridine, 198 inosine, and 1,043 Nm [7 Am, 255 Cm, 374 Gm, and 407 Um]). Finally, 51 AD-associated genes harbored 1,481 sites (468 m^6^A, 354 m^5^C, 296 pseudouridine, 102 inosine, and 261 Nm [74 Cm, 84 Gm, and 103 Um]); notably, no Am sites were detected in these genes.

We further quantified the co-occurrence within the modification types based on the number of genes containing 1 to 10+ unique modification sites (**Fig. 2b**), where each modification type has similar but distinct distributions. For example, the Nm and inosine modifications have far fewer co-occurrences while m^6^A (3,589 genes), m^5^C (2,121 genes), and pseudouridine (1,752 genes) have more genes with 10+ sites than with any individual count from 1 to 9 (**Fig. 2b**).

We identified 51 AD-associated genes with ≥1 modification, including *MAPT, PSEN1, CLU, APP, BIN1, ABCA7*, and *APOE* (**Fig. 2c**). *CTSB, SORT1, ANKH, MAPT, ANK3*, and *PSEN1* had the most modifications that passed our criteria, while *APOE* was ranked 25^th^ based on total sites modified. The estimated number of modifications for each gene should be interpreted with caution, however, because estimates for longer genes and isoforms (e.g., *ABCA7*), or those with lower expression, are likely underrepresented because they may not have adequate coverage through the 5’ end in this study to reliably capture all modifications. For example, the RefSeq mRNA isoform for *ABCA7* (NM_019112) is 6,815 nucleotides, where zero reads covered the full isoform in this study (i.e., long with low expression; median CPM = 3.49), while *APOE* (NM_000041) is only 1,166 nucleotides with deep coverage throughout the gene (i.e., short with high expression; median CPM = 512.79).

We next examined which modification types were present within each of the 51 modified AD genes. m^6^A was the predominant modification in 29 of the 51 modified AD genes, reflecting its transcriptome-wide prevalence (35.5% of sites). For example, in *MAPT* (ENST00000351559), all five modification types were detected, including three of the four Nm (**Fig. 2d**). Within an approximately 250-nucleotide stretch of the 3’ UTR, we observed six modified sites spanning five modification types including m^6^A (chr17:46,027,538; 26% modified), inosine (chr17:46,027,644; 29%), Um (chr17:46,027,684; 21%), Cm (chr17:46,027,738; 18%), a second m^6^A (chr17:46,027,772; 57%), and pseudouridine (chr17:46,027,785; 29%).

In aggregate, the composition of sites across AD genes closely resembled the transcriptome-wide composition (m^6^A: 31.6% vs. 35.5%; m^5^C: 23.9% vs. 23.9%; pseudouridine: 20.0% vs. 20.7%; Nm: 17.6% vs. 16.6%) with the exception of inosine, which was approximately two-fold more frequent in AD genes (6.9% vs. 3.2%). Thus, apart from inosine, raw site counts within AD genes largely reflect overall modification prevalence. Because this aggregate similarity could mask patterns within individual genes, we hypothesized that specific RNA modification types are enriched in AD-associated genes relative to their transcriptome-wide distributions. To test this, we compared the proportion of each modification type within each modified AD gene to its proportion transcriptome wide. Relative to the transcriptome-wide composition of sites, 10 AD genes were significantly enriched for a single modification type (one-sided Fisher’s exact test, Benjamini–Hochberg false discovery rate (FDR) <0.05; **Fig. 2e**) such as m^6^A (*MAF* and *RASGEF1C*), inosine (*SPPL2A, CTSB*, and *WDR12*), pseudouridine (*APP* and *PLEKHA1*), and Cm (*SPI1, GRN*, and *SLC2A4RG*).

### Alzheimer’s disease-specific analyses

Beyond general characterization, we performed three AD-related analyses, testing for (1) differentially modified nucleotides between cases and controls transcriptome wide, (2) associations between mRNA modifications and the *APOE*ϵ4 allele (rmQTL) in the five *APOE*ϵ3/ϵ4 heterozygotes, and (3) pairwise co-occurrence associations between RNA modifications within *APOE*. For the second analysis, we only tested associations with *APOE*ϵ4 because none of the subjects included in this study carry the *APOE*ϵ2 allele. In samples with confirmed *APOE*ϵ3/ϵ4 genotypes, the T and C alleles at rs429358 identify RNA molecules originating from the ϵ3 and ϵ4 haplotypes, respectively.

#### Transcriptome-wide differential modification analysis

We identified three RNA modification sites that were preliminarily differentially modified between conditions based on the percentage of RNA molecules carrying the modification (FDR < 0.05), including one m^6^A, one pseudouridine, and one Um (**Fig. 2f**). These sites were located in *MSANTD1* (chr4:3,256,510; balanced map p = 6.38 × 10^−9^; adjusted p = 9.78 × 10^−4^; balanced effect size = −0.28), *ADCK1* (chr14:77,933,522; balanced map p = 6.12 × 10^−8^; adjusted p = 5.67 × 10^−3^; balanced effect size = 0.38), and *NDUFS1* (chr2:206,122,652; balanced map p = 8.81 × 10^−7^; adjusted p = 0.030; balanced effect size = −0.22), respectively. In total, we tested 470,973 sites that met our inclusion criteria of ≥10% modification rate with ≥30 reads of coverage in at least three cases and three controls. Of the 470,973 sites, 153,173 were m^6^A, 117,204 m^5^C, 92,646 pseudouridine, 16,753 inosine, 406 Am, 21,042 Cm, 33,518 Um, and 36,231 Gm. No *APOE* modifications differed significantly between cases and controls but based on results from our second set of analyses, we expect that some will associate in a larger cohort because of the associations we found with rs429358. *There were no significant m*^*5*^*C hits, which may be due to a known m*^*5*^*C-specific* processing bug *in modkit dmr (version 0*.*6*.*4), but this bug should not affect other results, and we will update results once a bug fix is released*.

#### Modifications associated with the *APOE*ϵ4 allele

During our analyses, we specifically investigated RNA modifications in *APOE* because of its critical role in AD and initially noticed several highly prevalent modifications, including two m^6^A modifications (chr19:44,908,670 & chr19:44,908,679) and one m^5^C (chr19:44,908,671) that were all within 14 nucleotides of rs429358. The m^5^C is adjacent to the first m^6^A. Based on their proximity to rs429358, we wanted to test whether they are associated with *APOE*ϵ4 compared to ϵ3; this analysis was limited to the five *APOE*ϵ3/ϵ4 heterozygotes. The average modification rate for the two m^6^A sites and the m^5^C were approximately 45.0%, 35.6%, and 32.2%, respectively, across all mRNA molecules (i.e., combined between mRNA molecules from both *APOE* alleles, within subject) for the *APOE*ϵ3/ϵ4 heterozygotes. Of the five heterozygotes included in this study, three were cases. We also observed an m^5^C on the rs429358-C allele, itself, for approximately 11.5% of mRNA molecules arising from the *APOE*ϵ4 allele across the five carriers (**Fig. 3a,b**). No modifications passed our thresholds for the rs429358-T allele or the allele for the rs7412-C allele in this cohort. Since our cohort did not include any *APOE*ϵ2 carriers, we cannot report for the rs7412-T allele.

**Figure 3.**
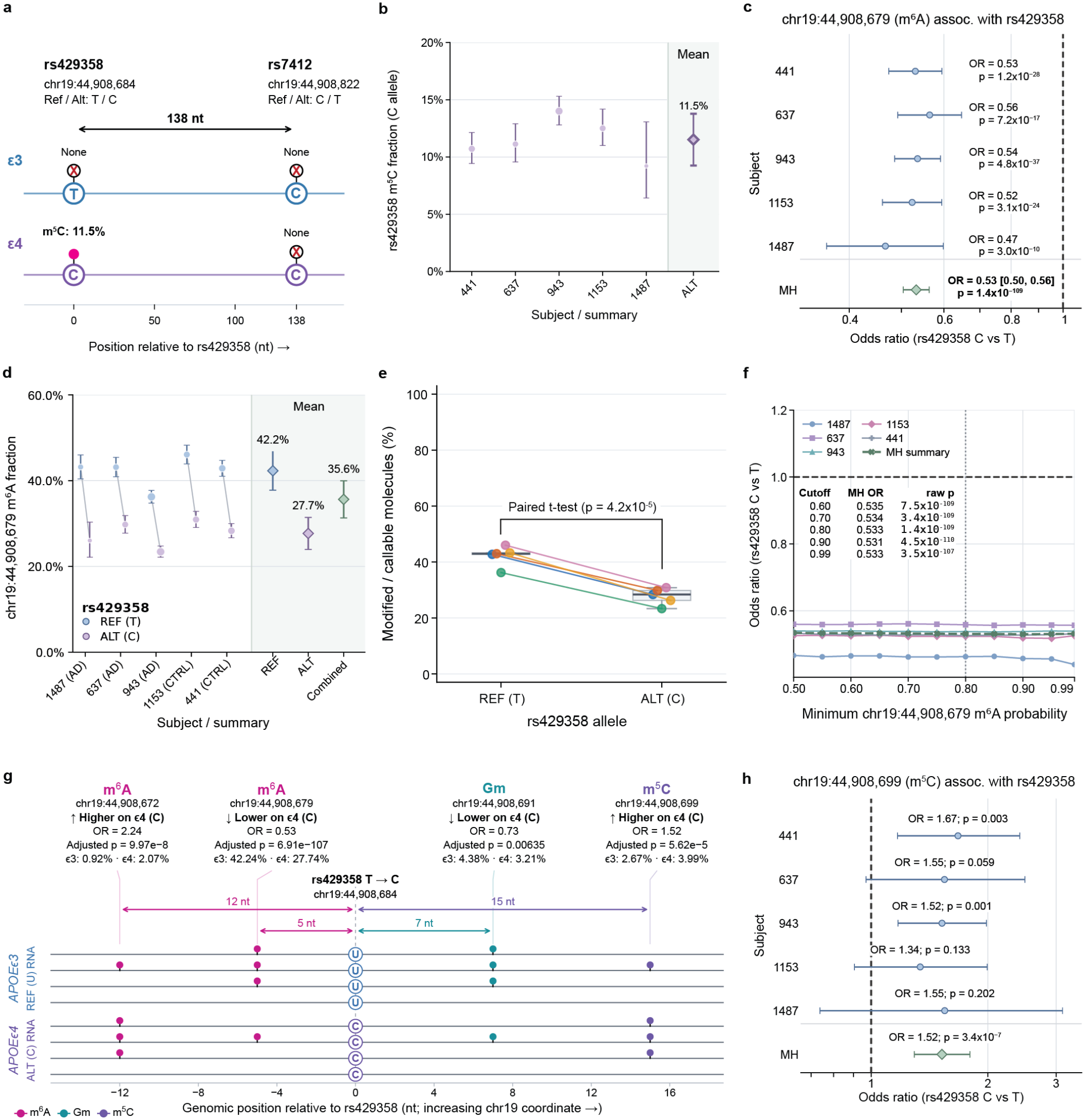
The rs429358-C allele is modified, and four mRNA modifications are associated with the *APOE*ϵ4 allele. We performed several AD-specific association tests for mRNA modifications in a cohort of six cases and six controls. (**a**) Schematic of rs429358 and rs7412 on the *APOE*ϵ3 and *APOE*ϵ4 haplotypes of the five *APOE*ϵ3/ϵ4 heterozygotes; the magenta circle marks the m5C detected on the rs429358-C allele (11.5% of ϵ4 molecules), and red X’s mark alleles with no modification passing our thresholds. ϵ2 and ϵ3r are not shown (absent from cohort). **(b**) We observed an m^5^C modification on 11.5% of mRNA molecules originating from the *APOE*ϵ4 haplotype (i.e., the C allele of rs429358) of five *APOE*ϵ3/ϵ4 heterozygotes. (**c**) Forest plot showing an m^6^A modification at chr19:44,908,679 is downregulated on mRNA molecules originating from the *APOE*ϵ4 haplotype across the five *APOE*ϵ3/ϵ4 heterozygotes (Mantel-Haenszel combined OR = 0.53 [0.50, 0.56]; Cochran-Mantel-Haenszel p = 1.4 × 10^−109^). (**d**) The m^6^A modification at chr19:44,908,679 is downregulated by 14.5 percentage points, on average across the five heterozygotes. (**e**) To further assess whether the effect was consistent across all heterozygotes, we performed a paired t-test, treating each subject as the independent observations, which was also significant (p = 4.19 × 10^−05^). (**f**) Because analyses require setting thresholds to determine whether a given nucleotide is canonical or modified, we assessed whether the association between the m^6^A modification at chr19:44,908,679 and rs429358 is robust across modification probability thresholds between 0.50 and 0.99. The association was consistent across the range. (**g**) An illustration showing all four modifications associated with rs429358 that are within 15nt of the SNP; two were upregulated on the ϵ4 haplotype and two were downregulated. The “lollipops” represent a modification at that position. (**h**) Forest plot showing an m^5^C located at chr19:44,908,699 is upregulated on mRNA molecules originating from the ϵ4 haplotype. Scripts used to create figures were generated by ChatGPT and human verified.

We tested for association between the three modifications described and the *APOE*ϵ4 allele in the five *APOE*ϵ3/ϵ4 heterozygotes, where we first used a Fisher’s exact test within each individual and then calculated a Mantel-Haenszel (MH) combined odds ratio (OR)^87^ and a Cochran-Mantel-Haenszel (CMH) p-value^88^ across the subjects. The m^6^A modification located at chr19:44,908,679 was strongly associated with the *APOE*ϵ4 allele (MH combined OR = 0.53 [0.50, 0.56]; CMH p = 1.4 × 10^−109^; **Fig. 3c**). Plotting the percent modified for each *APOE* allele shows the m^6^A is clearly and consistently downregulated on RNA molecules arising from the ϵ4 allele (27.7%) compared to the ϵ3 allele (42.2%)—a difference of 14.5 percentage points (**Fig. 3d**). There is no obvious difference in percent modified molecules between cases and controls in these five heterozygotes (**Fig. 3d**). To formally test whether the effect was consistent across all heterozygotes, we performed a paired t-test, treating each subject as the independent observations (rather than the reads), which was also significant (p = 4.19 × 10^−5^; **Fig. 3e**); this test assumes normality, however, which our current sample size cannot confirm. To test the robustness of this association, we assessed whether our selected confidence thresholds for what constitutes a modified nucleotide (0.80) affects the association by re-testing the association at probability thresholds between 0.5 and 0.99 (**Fig. 3f**); the association was robust at all thresholds.

Because *APOE* had deep coverage, we performed a more comprehensive analysis to quantify its rarer mRNA modifications and test which are associated with the ϵ4 allele. Thus, we identified all modifications with rates of 1%, 5%, 10%, 20%, and 30%, where we observed 502, 73, 25, 7, and 5, respectively, demonstrating a wide range of estimates, depending on the minimum modification rate. At all five rates, we required that the modification be observed in at least six of the twelve subjects and have ≥1,000 reads of coverage. Despite having deep coverage through the entire gene, most of the observed modifications were in *APOE*’s final exon (exon 4), where, for example, 62 of 73 modifications (84.9%) with rates ≥5% were in exon 4.

We tested association between all modifications with ≥1% modification rate and the *APOE*ϵ4 allele. Most of the modifications at 1% are likely false positives, given the inherent error rate in nanopore sequencing of >1%, but modifications associated with the *APOE*ϵ4 allele should be considered for validation experiments. Of the 502 *APOE* modifications with ≥1% modification rate, four were associated with the *APOE*ϵ4 allele (including the m^6^A modification located at chr19:44,908,679 previously described) having a Benjamini–Hochberg adjusted p-value <0.05—all within 15 nucleotides of rs429358 (**Fig. 3g**). The three additional modifications were an m^6^A located at chr19:44,908,672 (MH combined OR [95% CI] = 2.24 [1.72, 2.94]; adjusted CMH p = 4.0 × 10^−10^, a Gm located at chr19:44,908,691 (MH combined OR [95% CI] = 0.73 [0.63, 0.85]; adjusted CMH p = 5.1 × 10^−5^, and an m^5^C located at chr19:44,908,699 (MH combined OR [95% CI] = 1.52 [1.29, 1.80]; adjusted CMH p = 3.4 × 10^−7^; **Fig. 3h**). However, in ONT’s nanopore sequencing, up to approximately five consecutive nucleotides contribute to the measured ionic current^89–91^, allowing RNA modifications to perturb signals at adjacent positions and potentially cause false-positive modification calls^92–94^. The m^6^A located at chr19:44,908,672 is both rare (1.4% modification rate) and is only two nucleotides downstream from the m^6^A located at chr19:44,908,670 (45.0% modification rate) and the alleged m^5^C located at chr19:44,908,671. Being both rare and adjacent to other modifications could suggest it is a false positive.

Because dRNAseq is a newer approach, independent validation is essential. We validated the two m^6^A modifications located at chr19:44,908,670 and chr19:44,908,679 using GLORI-qPCR performed by a commercial service provider (CD Genomics, Shirley, NY, USA). The dRNAseq data estimated these modifications were present at 45.0% and 35.6%, respectively, while GLORI-qPCR estimated them at 49.5% and 7.0%, respectively. The discrepancy for the latter may be due to selected confidence thresholds in our current analysis or because of qPCR bias. In either case, both modifications validated, but further work will be needed to assess the true rate for the second m^6^A. We will validate additional modifications in the future.

#### Modification-modification associations within *APOE*

After testing associations between the RNA modifications and rs429358, we then tested for associations between all possible pairs of the 73 modifications with rates ≥5%. We used either an exact conditional test of independence for sample-stratified 2 × 2 tables or a Cochran–Mantel–Haenszel (CMH) chi-squared test without continuity correction when exact computation exceeded predefined limits and approximation safeguards were satisfied. Between the 73 modifications included in this analysis, we identified 344 significant modification-modification associations with an adjusted p-value <0.05 (**Fig. 4a,b**), where 295 were between modifications exclusive to exon 4 and all but six pairs had at least one of the modifications in exon 4. There were no significant associations with an inosine. Of the 344 associations, twelve were between modifications directly adjacent to each other, all of which had odds ratios >1, which could suggest that at least one of the two modifications in each pair is a false positive resulting from signal interference when identifying modifications, as described. Odds ratios <1 may be less likely to result from a false positive.

**Figure 4.**
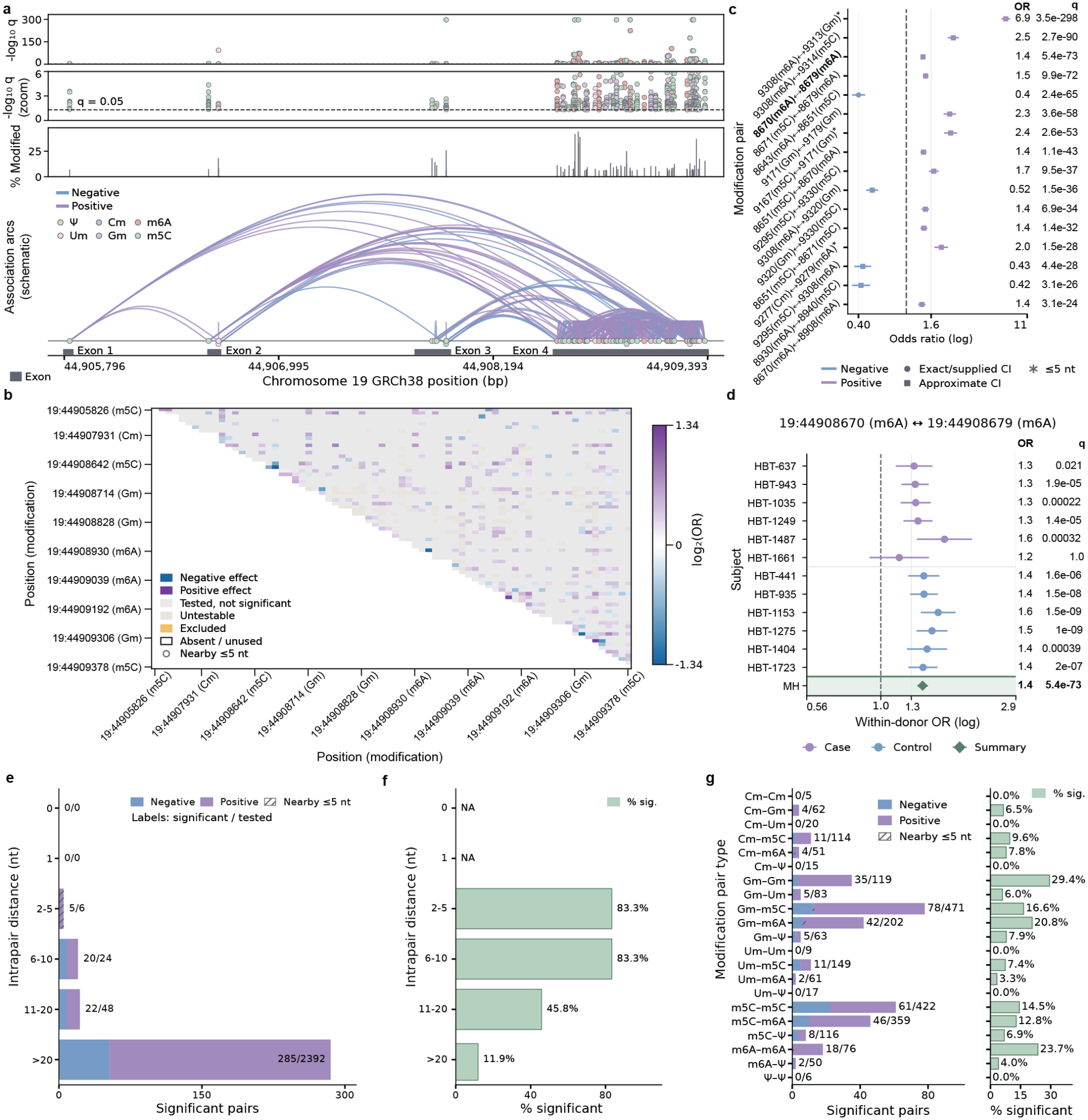
There were 344 modification-modification associations within *APOE*. (**a**) We mapped the 344 significant modification-modification associations throughout *APOE* arising from 73 distinct mRNA modifications with rates ≥5%. Of the 73 modifications, 62 (84.9%) were within exon 4 even though *APOE* had deep coverage throughout the gene in this experiment (>1,000 reads throughout in all twelve samples). Benjamini-Hochberg-adjusted p-values ranged from 3.54e-298 to 0.05, shown as a Manhattan plot on top with a zoomed in version below. We also show the mean percent of modified RNA molecules for all 73 modifications included here. (**b**) A heatmap showing the log_2_ of the odds ratio (OR) for all significant pairwise comparisons, ignoring those that are ≤5 nucleotides from each other. (**c**) A forest plot showing the top 16 modification-modification associations, sorted by adjusted p-value. The bolded association is between the two most common m^6^A mRNA modifications, including the one at location chr19:44,908,679 that is downregulated on the *APOE*ϵ4 allele. (**d**) A forest plot for the association between chr19:44,908,670 and chr19:44,908,670 showing individual samples (by disease status) and the Mantel–Haenszel summary OR. (**e**) The number of significant associations by distance between modifications. (**f**) The percentage of significant associations for each group in **e**. (**g**) The number of significant associations for each type of modification and their percentages. Scripts used to create figures were generated by ChatGPT and human verified.

Ignoring all significant pairs within ≤1 nucleotide from each other, odds ratios for the top 16 (by p-value) ranged from 0.4 to 6.9 (p-values ranged from 3.1 × 10^−24^ to 3.5 × 10^−298^; **Fig. 4c**). Among the top 15 was a positive association between the two most prevalent m^6^A modifications, mentioned previously (chr19:44,908,670 & chr19:44,908,679; OR = 1.4; adjusted p = 5.4 × 10^−73^; **Fig. 4c,d**), but without a statistical difference between cases and controls (adjusted p = 0.99); the latter modification was downregulated on the *APOE*ϵ4 allele (**Fig. 3c,d**), but the former was not associated with the *APOE*ϵ4 allele, suggesting a more complicated relationship between these features.

We then assessed how distance between pairs affected associations and observed that, while most significant associations were between modifications >20 nucleotides apart (**Fig. 4e**), the percentage of significant associations decreased as distance increased (**Fig. 4f**). Whether this trend is due to technical or biological reasons is unclear and will require significant additional work and experiments.

We then assessed which combinations of modification types had the most significant associations and the greatest percentage of which were significant (**Fig. 4g**). The most common were Gm-m^5^C (78 significant associations out of 441 tested pairs; 16.6%), followed by m^5^C-m^5^C pairs (61 of 422; 14.5%), followed by m^5^C-m^6^A (46 of 359; 12.8%), and Gm-m^6^A (42 of 202; 20.8%).

## Discussion

Here, we performed a proof-of-principle study to assess mRNA modifications across the transcriptome in human brain from six AD cases and six sex- and agematched controls, similar to recent work by Byrne et al.^72^ Our unique and most compelling results, however, were those related to modifications to *APOE* mRNA. Specifically, we identified four modifications that were either up or down regulated on RNA molecules originating from the *APOE*ϵ4 haplotype (relative to ϵ3) among *APOE*ϵ3/ϵ4 heterozygotes. We also identified 344 associations between 73 unique *APOE* modifications having ≥5% modification rate (modification-modification associations). *APOE* is the top known genetic risk gene for AD, and RNA modifications are an understudied area across biology, including in AD. These analyses and associations—made possible by the nucleotide-level, single-molecule resolution afforded by ONT’s direct RNA sequencing (dRNAseq)—open an entirely new area of research in AD biology, including by potentially providing critical insight into *APOE*’s involvement in AD development and progression.

While working with this unique data, one of the biggest open questions we identified was regarding the ideal confidence thresholds for determining whether a given nucleotide was modified or canonical—or whether there was insufficient evidence for either, requiring us to exclude the read for that given nucleotide. At face value, requiring a maximal confidence value for a modification seems like the obvious solution, but the thresholds also directly affect the number of reads that can be used for downstream analyses.

Similarly, because the probabilities between whether a given nucleotide is modified or canonical are directly linked (i.e., sum to 1), using an extremely strict threshold may be excessive. For example, with a modification threshold of 0.99, researchers must determine how to treat any read with a probability <0.99 (e.g., 0.98), where the two simplest solutions are to (1) treat it as a canonical nucleotide (likely incorrect); or (2) exclude the read from downstream analyses (also likely incorrect). The same challenge exists when selecting a threshold for accepting whether a nucleotide is canonical.

Another question remains for whether the two thresholds (i.e., for canonical or modified nucleotides) should always be identical. For example, if 0.99 were selected for one threshold and 0.8 were selected for the other, then reads where either value is between 0.8 and 0.99 will fall into the same buckets described above. Studies by both Byrne et al.^72^ and Hewel et al.^95^ have investigated the impact of these thresholds, but the ideal solution remains unclear to us. In this study, we used 0.8 for both thresholds.

We validated two *APOE* m^6^A modifications (chr19:44,908,670 & chr19:44,908,679) using GLORI-qPCR among the 73 *APOE* modifications we identified with ≥5% modification rate. The m^6^A located at chr19:44,908,679 was our strongest statistical association (MH combined OR = 0.53 [0.50, 0.56]; CMH p = 1.4 × 10^−109^), being downregulated on mRNA molecules originating from the *APOE*ϵ4 haplotype with a difference of 14.5 percentage points (ϵ3: 42.2%; ϵ4: 27.7%). Although this association is both consistent and strong among the five ϵ3/ϵ4 heterozygotes included in this study when treating each subject as independent observations (p = 4.19 × 10^−5^), confirming these results in a larger cohort is necessary and is the biggest limitation of the current version of this study. We are currently testing this in a larger cohort.

For reference, the sequence motif for the m^6^A located at chr19:44,908,679 was GGACG, which does not perfectly match the canonical D (A, G, or U), R (A or G), A, C, H (A, C, or U; DRACH) motif associated with m^6^A modifications (i.e., the final nucleotide should not be a G). Notably, previous studies have demonstrated METTL3-dependent methylation at non-canonical GGACG motifs^96^ and identified methylated GGACG among sequences enriched *in vitro* binding selections with the m6A reads YTHDC1, YTHDF1, and YTHDF2^97^. This suggests that the noncanonical sequence context does not necessarily preclude m^6^A deposition or reader recognition, although the regulatory protein acting at this specific *APOE* site remains unknown.

We also identified 344 associations between modifications that suggest a complex network of carefully choreographed biological signals within *APOE* alone. A larger cohort will be critical to clarifying which are genuine and which combinations could contribute to *APOE*’s AD etiology.

If we assume that the primary associations reported herein are real and reproducible, the next critical question is whether these modifications play a direct role in AD development and progression, which will require larger studies to discern whether, for example, the m^6^A located at chr19:44,908,679 is associated with disease independently of the *APOE* haplotype (i.e., ϵ2, ϵ3, or ϵ4) and what effect, if any, it has on the protein (e.g., abundance).

Being independently associated with disease would suggest that this mRNA modification biologically modifies disease through a molecular mechanism that is independent of the *APOE* haplotype. Even if it is not independently associated with disease, however, it could still be directly involved and essential to *APOE*’s AD mechanism. Thus, regardless of whether the m^6^A located at chr19:44,908,679 is independently associated with disease, extensive experimental work will be essential to understand how it and other mRNA modifications in *APOE* (and other AD genes) are involved in disease.

If the modification located at chr19:44,908,679 is ultimately involved in disease, understanding the exact mechanism will be critical. mRNA modifications are understudied, but previous work suggests that a given modification type can have unique downstream consequences, based on context^98–100^. Thus, what effect this modification could have on downstream proteins is currently unclear.

Given this strong association, and that RNA modifications are critical biological signals, it is likely that this or other RNA modifications are directly involved in disease. Additionally, even if this specific *APOE* modification is not directly involved in disease, it and other modifications could potentially be targeted as a biomarker for improving pre-symptomatic disease diagnosis or to selectively down- or up-regulate mRNA molecules (e.g., those originating from a given *APOE* haplotype), thereby reducing risk for disease early in life.

A larger study could also assess whether individual mRNA modifications associate with neuropathological variables, similar to that performed by Shade et al.^6^ Thus, further investigation into mRNA modifications in *APOE* and other AD genes is an invaluable new direction in AD research.

## Methods

### PubMed search

We estimated the number of publications related to *APOE* and AD by searching PubMed with the following search query: (APOE OR “apolipoprotein E”) AND (Alzheimer OR Alzheimer’s). This search returned exactly 16,917 publications, as of October 15, 2026.

### Sample collection, RNA isolation, and quality control

Frozen postmortem human dorsolateral prefrontal cortex (Brodmann area 9/46) specimens from twelve samples (six Alzheimer’s disease cases and six non-demented age-sex-matched controls) that were obtained from the Arizona Study of Aging and Neurodegenerative Disorders and Brain and Body Donation Program at Banner Sun Health Research Institute (Banner)^101^ tissue was stored at −80ºC until processing. All samples self-reported their race as White, with eight who selfreported Hispanic ethnicity (n=4 AD; n=4 controls); the study was limited to White participants due to insufficient representation of other groups. All samples were between the ages of 68 and 89 years old at their time of death. All samples had a reported postmortem interval time, defined as the time from death to autopsy, of less than 5 hours. Cohort demographics are summarized in **Table 1**.

*Exactly 24 additional samples have been collected from the University of Kentucky’s Sanders-Brown Center on Aging brain bank that are currently being processed as follow-up samples for this study. Their demographics will be reported in a future version of this pre-print*.

Processing began with approximately 20mg of cortical gray matter tissue fragmented on dry ice, transferred to a pre-cooled 1.5 mL low-binding Eppendorf tubes (Eppendorf, cat. no. 022431021). Samples were processed in four batches of twelve RNA isolations equating to 48 total samples, with four technical replicates per sample, with each batch balanced for Alzheimer’s disease cases and controls to minimize batch effects. Total RNA was isolated using the Qiagen RNeasy Lipid Tissue Mini RNA Extraction kit (Qiagen, cat. No. 74804) according to protocol v.HB-0144-004. Briefly, specimen aliquots were kept on dry ice until lysis in 1.5 mL low-binding Eppendorf tubes. Specimens were homogenized for approximately 2 minutes using a plastic pestle (Kontes Pellet Pestle, VWR, cat. no. KT749521-1500) in 1mL of QIAZol lysis reagent. Samples remained on ice to maintain RNA integrity while other samples were homogenized. Samples were then placed at room temperature (15-25ºC) for 5 minutes followed by adding 200 μL chloroform and inverting the tube for 15 seconds. The lysates were placed on the benchtop at room temperature for 2 minutes before centrifugation at 12,000*g* for 15 minutes at 4ºC. Following centrifugation approximately 600 µl of the upper aqueous phase was transferred to a fresh 2 mL lowbinding Eppendorf tube. Total RNA was precipitated by adding 1 times volume of 70% ethanol and then applied to a RNeasy Mini spin column for centrifugation for 15 seconds at 8,000*g* at room temperature (flow-through was discarded). The column was then washed twice 700 μL of Buffer RW1, centrifuged for 15 seconds at 8,000*g* each time (flow-through was discarded). Then, 500 μL of Buffer RPE was added to the column and centrifuged for 15 seconds at 8,000*g* (flow-through was discarded). Following washing the column was transferred to a fresh 1.5 mL low-bind collection tube and the RNA was eluted by the addition of 30 μL of Elution Buffer, followed by 1 minute centrifugation at 8,000*g*. Eluted RNA was immediately placed on ice to minimize RNA degradation and proceeded to quality control analysis. RNA quality was determined initially by the NanoDrop spectrophotometer (A260:A280 and A260:A230 absorbance ratios) and then via Agilent Fragment Analyzer 5200 with the RNA (15 nt) DNF-471 kit (Agilent). All specimen used for sequencing had nanodrop ratios >1.8 and RNA quality numbers >8.0 on the fragment analyzer.

### Library preparation, sequencing, and base calling

Isolated RNA was kept on ice until quality control testing was completed as described above. Long-read dRNAseq library preparation commenced, utilizing the Oxford Nanopore Technologies dRNAseq kit (SQK-RNA004). The protocol was performed according to the manufacture’s specifications.

Library preparation required 1-1.3ug of total RNA per sample as input. Poly(A) enrichment is inherent to this protocol. Each sample was sequenced on two FLO-PRORNA004 flow cells on the Prome-thION P24 platform, with one library loaded per flow cell and each run lasting 72 hours. Data were acquired on a PromethION P24 compute tower with NVIDIA H100 GPUs using MinKNOW v24.06.14. Data was then base called with dorado^102^ (v2.1.2) with the base model as rna004_sup@v6.0.0 and used rna004_sup@v6.0.0_inosine_m6A_2OmeA@v1, rna004_sup@v6.0.0_m5C_2OmeC@v1, rna004_sup@v6.0.0_pseU_2OmeU@v1, and rna004_sup@v6.0.0_2OmeG@v1 for modification calls.

### Read preprocessing, genomic alignment, and quality control

Reads from dRNAseq were preprocessed with the quality score >7. The preprocessed reads were then aligned to the GRCh38 human reference genome using dorado alignment which uses a minimap^103^ wrapper (v2.1.2), using heuristics -x splice -k 14. Aligned reads were filtered for forward strand, primary alignments with MAPQ ≥10 were included with the resulting BAM alignment files sorted by genomic coordinates and indexed using SAMtools^104^ v1.24 before downstream analysis. Quality control reports and statistics were generated using Nanocomp^105^ v1.25.6.

### Medically relevant genes

Because clinicians typically interpret a genetic mutation in the context of a single isoform, which may not be expressed in the relevant tissue or cell type, knowing which tissues and cell types express each isoforms and modifications are essential for interpreting mutations in health and disease. To assess isoform expression and RNA modifications of these genes in the frontal cortex, we used a previously published list of medically relevant genes^85^, supplemented with genes linked to brain-related diseases^86^.

### RNA isoform quantification

Filtered BAM files were used for transcript identification and quantification using Bambu with Ensembl10 v.110 GTF annotation. Bambu was run in two steps within our Nextflow pipeline^86,106^ v23.10.0. Each sample’s BAM file was first preprocessed individually to generate an RDS file, and the RDS files from all twelve samples were then combined for joint transcript quantification across the cohort. Read-to-transcript assignments were retained (track_reads).

Bambu also generates a gene-level counts matrix that includes reads derived from both intronic and exonic regions. To generate an exon-derived gene-level expression matrix, we summed transcript-level counts across all transcripts assigned to each gene. Unless stated otherwise, gene-level expression values reported were derived from this exon-based count matrix.

For analyses quantifying gene bodies were defined as gene bodies with total counts per million (CPM) >1 in at least three samples. When quantifying gene bodies with multiple RNA isoforms, gene bodies were defined as their corresponding RNA isoforms were expressed at transcript-level total CPM >1 in at least three samples. For quantifying high-confidence RNA isoforms, RNA isoforms were defined as those expressed at a transcript-level total CPM >1 in at least three samples. This CPM > 1 cutoff follows our previous long-read RNA-seq study of aged human frontal cortex^86^, where it yielded orthogonally validated high-confidence isoforms, and has since been applied to DRS of AD frontal cortex^72^. Requiring CPM > 1 in at least three of the twelve samples increases confidence that retained genes and isoforms are reproducibly expressed across samples rather than driven by a single individual.

### RNA modification calling and per-site quantifica-tion

Per-site modification calls were summarized from the aligned BAM files using modkit pileup (v0.6.4), with technical replicates merged into one BAM file per sample. Five modifications were analyzed: m^6^A, inosine, and 2′-O-methyladenosine (Am) on adenosine; m^5^C and 2′-O-methylcytidine (Cm) on cytidine; pseudouridine and 2′-O-methyluridine (Um) on uridine; and 2′-O-methylguanosine (Gm) on guanosine. Calls within the first 4 and last 8 nucleotides of each read were excluded (--edge-filter 4,8). Calls were retained only if their probability was ≥0.8, applied to both canonical (unmodified) calls for each base (--filter-threshold) and modified calls for each modification (--mod-threshold). The maximum read depth considered per position was set to 60,000 (--max-depth=60000). Because several modifications are called on the same canonical base (e.g., m^6^A, inosine, and Am on adenosine), a separate pileup was run for each modification and restricted to that modification (--modified-bases). Pileups were output as bgzip-compressed, tabix-indexed bedMethyl files and combined across the twelve samples.

### Transcriptome-wide candidate site filtering

Following per-site quantification, candidate modification sites, defined by chromosome, position, and modification type, were retained when at least six of the twelve samples had a valid coverage of ≥30 reads with ≥10% of those reads called as modified. Requiring support from at least six samples ensures that a site is reproducibly modified rather than present by a single sample. Sites were then assigned to genes using the bambu-extended annotation. Intergenic sites or sites in overlapping genes were excluded so that each site mapped to a single gene. Remaining sites were retained if their gene was expressed at ≥1 count per million (CPM) in at least six of twelve samples.

To identify modification types enriched within individual AD-associated genes, we performed one-sided Fisher’s exact tests comparing the proportion of sites belonging to each modification type within each gene against the corresponding proportion across all other genes. Fold enrichment was calculated as the proportion of sites belonging to a given modification type within a gene divided by its proportion among all retained sites transcriptome wide. P-values were adjusted using the Benjamini–Hochberg method across all ADrelevant gene modification type comparisons, with FDR <0.05 considered significant.

### Differential modification analysis

Prior to testing, candidate sites were further restricted to those with ≥10% modification in at least three AD or at least three control samples, retaining sites modified in only one group. AD and control samples were then compared at these sites, separately for each modification, using modkit dmr pair with the six samples per group as replicates; at each site, only samples with ≥30 valid reads contributed to the test. After testing, sites were retained only if at least three AD and at least three control samples met this coverage threshold of ≥30 valid reads, including samples with no modified calls. This ensures that a site appearing unmodified in one group reflects a true absence of modification rather than insufficient coverage, and that both groups have adequate reads of sufficient coverage. Modkit calculates a balanced maximum a posteriori (MAP)-based p*-*value by comparing the posterior density of no difference in modification frequency between conditions with that of the observed difference, while balancing contributions across biological replicates. These values (balanced_map_pvalue) were adjusted using the Benjamini–Hochberg procedure separately for each modification type, and sites with a FDR <0.05 were considered differentially modified. Effect size was the Modkit balanced effect size, defined as the difference in the fraction of modified calls (control minus AD); positive values indicate higher modification in controls and negative values indicate higher modification in AD (e.g., 0.05 indicates 5 percentage points higher modification in controls). Volcano plots highlight sites that had an absolute effect size ≥0.05.

### RNA variant-modification associations (i.e., rmQTL) identification on *APOE*

To identify significant associations between rs429358 and RNA modifications within *APOE*, we analyzed RNA molecules spanning the SNP and each modification site in five samples with the *APOE*ϵ3/ϵ4 genotype. For each sample, molecules were classified by rs429358 allele (ϵ3: T; ϵ4: C) and target modification status to generate a 2 × 2 table. Molecules classified as canonical or carrying another modification were considered negative for the target modification. Unknown modification states were excluded. Modifications at the SNP coordinate were reported as modification frequencies on the eligible allele and were not included in REF–ALT association tests.

We used an exact conditional test of independence for sample-stratified tables (stratified_exact)^107^ when there were ≤2,001 possible total-overlap values and ≤2,000,000 estimated calculation operations. For each sample, allele and modification totals were held fixed, and the probability distribution of their overlap under independence was calculated. These distributions were combined across samples to obtain an exact, two-sided conditional p-value. Inference required at least three samples with all row and column marginal counts ≥3 and aggregate marginal counts ≥10 across informative samples. Samples without variation in either allele or modification status did not contribute to the stratified test. When exact calculation exceeded the configured computational limits, we used the Cochran–Mantel– Haenszel chi-squared test (cmh_uncorrected)^87,88^ without continuity correction, provided that every expected cell count in every informative sample was ≥5 and the total conditional variance was ≥10. Otherwise, inference was withheld. Both methods reported a Mantel– Haenszel common odds ratio comparing the odds of modification on ALT versus REF molecules.

To assess consistency across samples, we calculated the ALT−REF difference in modification proportions for each sample with callable coverage for both alleles. We calculated the mean difference, standard deviation, and 95% t confidence interval, weighting samples equally regardless of sequencing depth. We used a two-sided paired t-test to determine whether the mean difference differed from zero when at least three samples were available and the between-sample variance was nonzero. Confidence intervals required at least two samples and nonzero variance. The paired test treated samples as independent biological replicates and assumed approximately normally distributed sample differences.

We also calculated the proportion of samples whose modification difference agreed with the direction of the Mantel–Haenszel odds ratio. We used an exact, two-sided sign test to evaluate directional consistency, excluding samples with zero differences. To assess sensitivity to individual samples, we repeated the samplelevel analysis after removing each sample and reported the ranges of mean differences and estimable paired t-test p-values. Benjamini–Hochberg correction^108^ was applied separately to stratified association, paired t-test, and sign-test p-values. For stratified associations, unavailable tests were retained as p = 1 during adjustment but were reported as unavailable. For sample-level tests, correction included all pairs with an estimable corresponding test, regardless of stratified-test significance. Adjusted p-values <0.05 were considered significant within each test family. Analyses were implemented in Python using custom code with SciPy, NumPy, and statsmodels (see Code Availability).

### RNA modification-modification associations within *APOE*

To identify significant modification-modification associations within *APOE*, we used an exact conditional test of independence for subject-stratified 2 × 2 tables (stratified_exact)^107^ if there were less than 2,000 possible total-overlap values and less than 2,000,000 estimated calculation operations. Otherwise, we used the Cochran–Mantel–Haenszel chi-squared test (cmh_uncorrected)^87,88^ without continuity correction if approximation safeguards were satisfied. If safeguards were not satisfied, no test was performed. Associations with a Benjamini-Hochberg^108^ adjusted p-value <0.05 were considered significant.

For the exact conditional test of independence, associations were evaluated within samples while accounting for differences in each sample’s modification frequencies. For each sample, the numbers of molecules positive for modifications A and B were held fixed, and the probability distribution of their overlap under independence was calculated. These distributions were combined across samples to obtain an exact, two-sided conditional p-value. The calculation was implemented in Python using custom code with SciPy and NumPy.

When exact calculation exceeded the configured computational limits, we used the Cochran–Mantel– Haenszel approximation without continuity correction, provided that every expected cell count in every informative sample was at least 5 and the total conditional variance was at least 10. Otherwise, inference was withheld. Both methods reported a Mantel–Haenszel common odds ratio. Samples without variation in either modification did not contribute to the stratified test.

### GLORI-qPCR validation of m^6^A modifications within *APOE*

To validate m^6^A modifications at chr19:44908670 and chr19:44908679 within *APOE*, we used GLORI treatment followed by quantitative realtime PCR (GLORI-qPCR; performed by CD Genomics in Shirley, NY, USA). RNA samples from all twelve subjects were submitted for analysis. RNA concentration and purity were measured using a NanoDrop ND-1000 spectrophotometer. RNA integrity was assessed by denaturing agarose gel electrophoresis. The twelve samples were then pooled to create three samples to ensure sufficient RNA for the protocol.

RNA was fragmented at 94°C for 4 min and processed using the GenSeq m^6^A GLORI Kit (Cayman Chemical). Fragmented RNA was incubated in a 50-µL reaction containing glyoxal, dimethyl sulfoxide, sodium nitrite, saturated boric acid solution, and MES buffer (pH 6.0) at 50°C for 10 min. RNA was recovered by ethanol precipitation, resuspended in deprotection buffer containing 2 mM EDTA, and incubated at 95°C for 10 min. RNA was then purified by a second ethanol precipitation.

For cDNA synthesis, 0.8 µg RNA was combined with 0.5 µg N6 primers and 1.6 µL of 2.5 mM dNTP mix in a volume of 14.5 µL. The mixture was incubated at 65°C for 5 min and immediately chilled on ice for 2 min. First-strand buffer, DTT, RNase inhibitor, and SuperScript III reverse transcriptase were added to a final reaction volume of 20 µL. Reactions were incubated at 37°C for 1 min, followed by 50°C for 60 min. Reverse transcriptase was inactivated at 70°C for 15 min.

Separate forward primers were used to distinguish m^6^A and non-m^6^A templates, with a common reverse primer for each target site (**Table 2**). qPCR was performed in 384-well plates using 10-µL reactions containing 5 µL of 2x master mix, 0.5 µL each of 10 µM forward and reverse primers, 2 µL cDNA, and water. Amplification consisted of an initial incubation at 95°C for 10 min, followed by 40 cycles of 95°C for 10 s and 60°C for 60 s. Fluorescence was collected during the 60°C step.

**Table 2.**
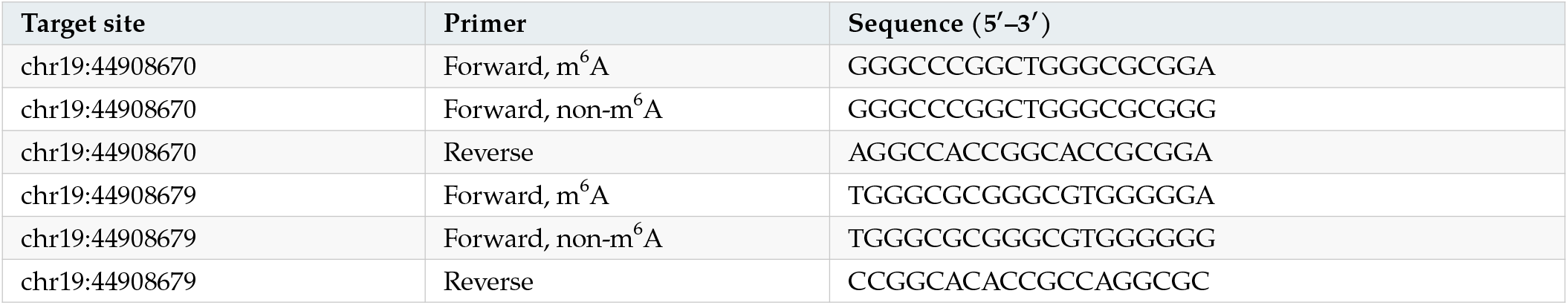
Primers used for GLORI-qPCR.

### Statistical analysis and figures and tables

Group comparisons for the demographic, neuropathological, tissue-quality, and sequencing characteristics used Welch’s t-test or the Mann–Whitney U test, as appropriate. Modification enrichment within each AD gene relative to the remainder of the transcriptome was quantified as log_2_ fold enrichment and tested using a one-sided Fisher’s exact test. Differential RNA modification was assessed with modkit dmr pair.

Figures and tables were generated using customized R (v4.5.2) scripts and customized Python (v3.10+) scripts. Data processing and manipulation performed with R used dplyr (v1.1.4), tidyr (v1.3.2) and data.table (v1.18.0). Data visualization with R was performed using ggplot2 (v4.0.1), CompexHeatmap (v2.24.1), EnhancedVolcano (v1.26.0), and circilze (v0.4.17). Data processing, manipulation, and visualization were performed using Python with pandas (v2.2.1), numpy (v1.26.4), seaborn (v0.13.2), re (v2.2.1), and polars (v1.39.3). See ‘Code Availability’ for access to the customized scripts used to generate figures and tables.

## Abbreviations

(m^6^A): N6-methyladenosine
(m^5^C): 5-methylcytosine
(m^1^A): N1-methyladenosine
(Ψ): pseudouridine
(A-to-I): Adenosine-to-Inosine
(ONT): Oxford Nanopore Technologies
(PCR): polymerase chain reaction
(cDNA): complementary DNA

## Code availability

The code will be made available in a future version of this preprint.

## Data availability

The data will be made available in a future version of this preprint.

## Use of Large Language Models

Large language models (LLMs) were utilized to help improve the clarity, conciseness, and flow of the manuscript text. Additionally, LLMs assisted in generating code for data analysis. All LLM generated text and code was reviewed by a human to ensure accuracy.

## Rigor and reproducibility

This study was conducted under the oversight of the University of Kentucky Institutional Review Board. Read preprocessing, alignment, quality control, and filtering were performed using custom scripts. Transcriptome quantification for the dRNAseq data were performed using a NextFlow^109^ (v.23.10.0) pipeline, which enables scalable and reproducible workflows with software containers. For most analyses, we employed Singularity/Apptainer^110^ (v.3.8.0-1.el8) containers. Instructions for accessing the containers are available in the project’s GitHub repository. Any deviations from standard manufacturer protocols are detailed in the **Methods**. The code will be made available in a future version of this preprint. The data will be made available in a future version of this preprint.

## Author contributions

Conceptualization, supervision, visualization, investigation, writing – original draft, reviewing and editing was done by G.A.F. and M.T.W.E. Brain tissue storage was managed by L.A.G., who also contributed to sequencing data generation. M.L.P., B.A.H., M.E.W., D.W.F, and L.A.J. provided important intellectual contributions. P.T.N., G.E.S., and T.G.B., provided the invaluable brain samples and pathology data. All authors have read and agreed to the published version of the manuscript.

## Acknowledgements

G.A.F. and M.T.W.E. are members of the Human RNome Project consortium. We appreciate the contributions of the Sanders-Brown Center on Aging at the University of Kentucky and The Brain and Body Donation program at Banner Sun Research Institute. We are deeply appreciative of the research participants and their families who made this research possible. We would like to thank the University of Kentucky Center for Computational Sciences and Information Technology Services Research Computing for their support and use of the Morgan Compute Cluster, Lipscomb Compute Cluster, and associated research computing resources. We thank CD Genomics (Shirley, NY, USA) for technical assistance validating m^6^A modifications via GLORI-qPCR. We thank the reviewers for their sincere and meaningful contributions to improving the quality of the manuscript.

## References

1. Strittmatter, W. J. et al. Apolipoprotein E: high-avidity binding to beta-amyloid and increased frequency of type 4 allele in late-onset familial Alzheimer disease. Proceedings of the National Academy of Sciences of the United States of America 90, 1977–1981 (1993).

2. Saunders, A. M. et al. Association of apolipoprotein E allele ϵ4 with late-onset familial and sporadic Alzheimer’s disease. Neurology 43, 1467 (1993).

3. Corder, E. H. et al. Gene dose of apolipoprotein E type 4 allele and the risk of Alzheimer’s disease in late onset families. Science (New York, N.Y.) 261, 921–923 (1993).

4. EADB et al. Consensus meta-analysis of genome-wide association studies for Alzheimer’s disease and related dementias. Nature Genetics 58, 1214–1225 (2026).

5. Bellenguez, C. et al. New insights into the genetic etiology of Alzheimer’s disease and related dementias. Nature Genetics 54, 412–436 (2022).

6. Shade, L. M. P. et al. GWAS of multiple neuropathology endophenotypes identifies new risk loci and provides insights into the genetic risk of dementia. Nature Genetics 56, 2407–2421 (2024).

7. Williams, D. M., Mukadam, N., Davies, N. M. & Anderson, E. L. Understanding apolipoprotein E as a predominant cause of Alzheimer’s disease. Brain awag395 (2026) doi:10.1093/brain/awag395.

8. Murrell, J. R. et al. The fourth apolipoprotein E haplotype found in the Yoruba of Ibadan. American Journal of Medical Genetics Part B: Neuropsychiatric Genetics 141B, 426–427 (2006).

9. Persico, A. M. et al. Enhanced APOE2 transmission rates in families with autistic probands. Psychiatric Genetics 14, 73–82 (2004).

10. Kemper, K. E. et al. Genetic influence on within-person longitudinal change in anthropometric traits in the UK Biobank. Nature Communications 15, 3776 (2024).

11. Williams, D. M. et al. The proportion of Alzheimer’s disease attributable to apolipoprotein E. Preprint at 10.1101/2023.11.16.23298475 (2023).

12. Gustavsson, A. et al. Global estimates on the number of persons across the Alzheimer’s disease continuum. Alzheimer’s & Dementia 19, 658–670 (2023).

13. Holtzman, D. M. et al. Apolipoprotein E isoform-dependent amyloid deposition and neuritic degeneration in a mouse model of Alzheimer’s disease. Proceedings of the National Academy of Sciences of the United States of America 97, 2892–2897 (2000).

14. Deane, R. et al. apoE isoform-specific disruption of amyloid beta peptide clearance from mouse brain. The Journal of Clinical Investigation 118, 4002–4013 (2008).

15. Castellano, J. M. et al. Human apoE isoforms differentially regulate brain amyloid-β peptide clearance. Science Translational Medicine 3, 89ra57 (2011).

16. Nelson, P. T. et al. APOE-ε2 and APOE-ε4 correlate with increased amyloid accumulation in cerebral vasculature. Journal of Neuropathology and Experimental Neurology 72, 708–715 (2013).

17. Abner, E. L. et al. Diffuse Amyloid-β Plaques, Neurofibrillary Tangles, and the Impact of APOE in Elderly Persons’ Brains Lacking Neuritic Amyloid Plaques. Journal of Alzheimer’s disease: JAD 64, 1307–1324 (2018).

18. Liao, F. et al. Targeting of nonlipidated, aggregated apoE with antibodies inhibits amyloid accumulation. The Journal of Clinical Investigation 128, 2144–2155 (2018).

19. Lee, S.-I. et al. APOE4-carrying human astrocytes oversupply cholesterol to promote neuronal lipid raft expansion and Aβ generation. Stem Cell Reports 16, 2128–2137 (2021).

20. Shi, Y. et al. ApoE4 markedly exacerbates tau-mediated neurodegeneration in a mouse model of tauopathy. Nature 549, 523–527 (2017).

21. Shi, Y. et al. Microglia drive APOE-dependent neurodegeneration in a tauopathy mouse model. The Journal of Experimental Medicine 216, 2546–2561 (2019).

22. Wang, C. et al. Selective removal of astrocytic APOE4 strongly protects against tau-mediated neurodegeneration and decreases synaptic phagocytosis by microglia. Neuron 109, 1657–1674.e7 (2021).

23. Gratuze, M. et al. APOE Antibody Inhibits Aβ-Associated Tau Seeding and Spreading in a Mouse Model. Annals of Neurology 91, 847–852 (2022).

24. Sienski, G. et al. APOE4 disrupts intracellular lipid homeostasis in human iPSC-derived glia. Science Translational Medicine 13, eaaz4564 (2021).

25. Blanchard, J. W. et al. APOE4 impairs myelination via cholesterol dysregulation in oligodendrocytes. Nature 611, 769–779 (2022).

26. Victor, M. B. et al. Lipid accumulation induced by APOE4 impairs microglial surveillance of neuronal-network activity. Cell Stem Cell 29, 1197–1212.e8 (2022).

27. Lee, S. et al. APOE modulates microglial immunometabolism in response to age, amyloid pathology, and inflammatory challenge. Cell Reports 42, 112196 (2023).

28. Friday, C. M. et al. APOE4 reshapes the lipid droplet proteome and modulates microglial inflammatory responses. Neurobiology of Disease 212, 106983 (2025).

29. Nolt, G. L. et al. Microglia-derived APOE2 improves remyelination even in the presence of endogenous APOE4. Journal of Neuroinflammation 23, 18 (2025).

30. Goulding, D. S. et al. APOE4 drives maladaptive heterogeneity and immunometabolic responses of astrocytes. Journal of Neuroinflammation 23, 89 (2026).

31. Hu, J., LaDu, M. J. & Van Eldik, L. J. Apolipoprotein E attenuates beta-amyloid-induced astrocyte activation. Journal of Neurochemistry 71, 1626–1634 (1998).

32. Krasemann, S. et al. The TREM2-APOE Pathway Drives the Transcriptional Phenotype of Dysfunctional Microglia in Neurodegenerative Diseases. Immunity 47, 566–581.e9 (2017).

33. Ulrich, J. D. et al. ApoE facilitates the microglial response to amyloid plaque pathology. The Journal of Experimental Medicine 215, 1047–1058 (2018).

34. Kloske, C. M. et al. Inflammatory Pathways Are Impaired in Alzheimer Disease and Differentially Associated With Apolipoprotein E Status. Journal of Neuropathology and Experimental Neurology 80, 922–932 (2021).

35. Seaks, C. E. et al. Apolipoprotein E ε4/4 genotype limits response to dietary induction of hyperhomocysteinemia and resulting inflammatory signaling. Journal of Cerebral Blood Flow and Metabolism: Official Journal of the International Society of Cerebral Blood Flow and Metabolism 42, 771–787 (2022).

36. Arnaud, L. et al. APOE4 drives inflammation in human astrocytes via TAGLN3 repression and NF-κB activation. Cell Reports 40, 111200 (2022).

37. Bell, R. D. et al. Apolipoprotein E controls cerebrovascular integrity via cyclophilin A. Nature 485, 512–516 (2012).

38. Halliday, M. R. et al. Accelerated pericyte degeneration and blood-brain barrier breakdown in apolipoprotein E4 carriers with Alzheimer’s disease. Journal of Cerebral Blood Flow and Metabolism: Official Journal of the International Society of Cerebral Blood Flow and Metabolism 36, 216–227 (2016).

39. Montagne, A. et al. APOE4 leads to blood-brain barrier dysfunction predicting cognitive decline. Nature 581, 71–76 (2020).

40. Levi, O., Jongen-Relo, A. L., Feldon, J. & Michaelson, D. M. Brain area- and isoform-specific inhibition of synaptic plasticity by apoE4. Journal of the Neurological Sciences 229–230, 241–248 (2005).

41. Qiao, F., Gao, X.-P., Yuan, L.Cai, H.-Y. & Qi, J.-S. Apolipoprotein E4 impairs in vivo hippocampal long-term synaptic plasticity by reducing the phosphorylation of CaMKIIα and CREB. Journal of Alzheimer’s disease: JAD 41, 1165–1176 (2014).

42. Lin, Y.-T. et al. APOE4 Causes Widespread Molecular and Cellular Alterations Associated with Alzheimer’s Disease Phenotypes in Human iPSC-Derived Brain Cell Types. Neuron 98, 1141–1154.e7 (2018).

43. Li, X. et al. Astrocytic ApoE reprograms neuronal cholesterol metabolism and histone-acetylation-mediated memory. Neuron 109, 957–970.e8 (2021).

44. Braak, H. & Braak, E. Neuropathological stageing of Alzheimer-related changes. Acta Neuropathologica 82, 239–259 (1991).

45. Braak, H., Alafuzoff, I., Arzberger, T., Kretzschmar, H. & Del Tredici, K. Staging of Alzheimer disease-associated neurofibrillary pathology using paraffin sections and immunocytochemistry. Acta Neuropathologica 112, 389–404 (2006).

46. Khajouei, E. et al. Phenome-wide association of APOE alleles in the All of Us Research Program. EBioMedicine 117, 105768 (2025).

47. Zhang, X. et al. Causal association of circulating cholesterol levels with dementia: a mendelian randomization metaanalysis. Translational Psychiatry 10, 145 (2020).

48. Cappannini, A. et al. MODOMICS: a database of RNA modifications and related information. 2023 update. Nucleic Acids Research 52, D239–D244 (2024).

49. Boileau, E. et al. Sci-ModoM: a quantitative database of transcriptome-wide high-throughput RNA modification sites. Nucleic Acids Research 53, D310–D317 (2025).

50. Hengesbach, M. et al. Toward standardized epitranscriptome analytics: an inter-laboratory comparison of mass spectrometric detection and quantification of modified ribonucleosides in human RNA. Nucleic Acids Research 53, gkaf895 (2025).

51. Chujo, T. & Tomizawa, K. Human transfer RNA modopathies: diseases caused by aberrations in transfer RNA modifications. The Febs Journal 288, 7096–7122 (2021).

52. Dominissini, D. et al. Topology of the human and mouse m6A RNA methylomes revealed by m6A-seq. Nature 485, 201–206 (2012).

53. Meyer, K. D. et al. Comprehensive Analysis of mRNA Methylation Reveals Enrichment in 3’ UTRs and Near Stop Codons. Cell 149, 1635–1646 (2012).

54. Li, X. et al. Transcriptome-wide mapping reveals reversible and dynamic N(1)-methyladenosine methylome. Nature Chemical Biology 12, 311–316 (2016).

55. Squires, J. E. et al. Widespread occurrence of 5-methylcytosine in human coding and non-coding RNA. Nucleic Acids Research 40, 5023–5033 (2012).

56. Arango, D. et al. Acetylation of Cytidine in mRNA Promotes Translation Efficiency. Cell 175, 1872–1886.e24 (2018).

57. Nishikura, K. A-to-I editing of coding and non-coding RNAs by ADARs. Nature Reviews. Molecular Cell Biology 17, 83–96 (2016).

58. Walkley, C. R. & Li, J. B. Rewriting the transcriptome: adenosine-to-inosine RNA editing by ADARs. Genome Biology 18, 205 (2017).

59. Hodges, P. E., Navaratnam, N., Greeve, J. C. & Scott, J. Site-specific creation of uridine from cytidine in apolipoprotein B mRNA editing. Nucleic Acids Research 19, 1197–1201 (1991).

60. Chester, A., Scott, J., Anant, S. & Navaratnam, N. RNA editing: cytidine to uridine conversion in apolipoprotein B mRNA. Biochimica et Biophysica Acta (BBA) - Gene Structure and Expression 1494, 1–13 (2000).

61. Carlile, T. M. et al. Pseudouridine profiling reveals regulated mRNA pseudouridylation in yeast and human cells. Nature 515, 143–146 (2014).

62. Schwartz, S. et al. Transcriptome-wide mapping reveals widespread dynamic-regulated pseudouridylation of ncRNA and mRNA. Cell 159, 148–162 (2014).

63. Finet, O. et al. Transcription-wide mapping of dihydrouridine reveals that mRNA dihydrouridylation is required for meiotic chromosome segregation. Molecular Cell 82, 404–419.e9 (2022).

64. Ju, C.-W. et al. Quantitative CRACI reveals transcriptome-wide distribution of RNA dihydrouridine at base resolution. Nature Communications 16, 8863 (2025).

65. Dai, Q. et al. Nm-seq maps 2’-O-methylation sites in human mRNA with base precision. Nature Methods 14, 695–698 (2017).

66. Chu, J.-M. et al. Existence of Internal N7-Methylguanosine Modification in mRNA Determined by Differential Enzyme Treatment Coupled with Mass Spectrometry Analysis. ACS chemical biology 13, 3243–3250 (2018).

67. Zhao, Z. et al. QKI shuttles internal m7G-modified transcripts into stress granules and modulates mRNA metabolism. Cell 186, 3208–3226.e27 (2023).

68. Gilbert, W. V. & Nachtergaele, S. mRNA Regulation by RNA Modifications. Annual review of biochemistry 92, 175–198 (2023).

69. Enroth, C. et al. Detection of internal N7-methylguanosine (m7G) RNA modifications by mutational profiling sequencing. Nucleic Acids Research 47, e126 (2019).

70. Wiener, D. & Schwartz, S. The epitranscriptome beyond m6A. Nature Reviews. Genetics 22, 119–131 (2021).

71. Warminski, M., Sikorski, P. J., Kowalska, J. & Jemielity, J. Applications of Phosphate Modification and Labeling to Study (m)RNA Caps. Topics in Current Chemistry (Cham) 375, 16 (2017).

72. Byrne, A. et al. Direct RNA Sequencing reveals epitranscriptomic regulation of brain cells and Alzheimer’s Disease pathology. Preprint at 10.64898/2026.05.18.724443 (2026).

73. Ayub, M. & Bayley, H. Individual RNA base recognition in immobilized oligonucleotides using a protein nanopore. Nano Letters 12, 5637–5643 (2012).

74. Garalde, D. R. et al. Highly parallel direct RNA sequencing on an array of nanopores. Nature Methods 15, 201–206 (2018).

75. Jenjaroenpun, P. et al. Complete genomic and transcriptional landscape analysis using third-generation sequencing: a case study of Saccharomyces cerevisiae CEN.PK113-7D. Nucleic Acids Research 46, e38 (2018).

76. Keller, M. W. et al. Direct RNA Sequencing of the Coding Complete Influenza A Virus Genome. Scientific Reports 8, 14408 (2018).

77. Depledge, D. P. et al. Direct RNA sequencing on nanopore arrays redefines the transcriptional complexity of a viral pathogen. Nature Communications 10, 754 (2019).

78. Smith, A. M., Jain, M., Mulroney, L., Garalde, D. R. & Akeson, M. Reading canonical and modified nucleobases in 16S ribosomal RNA using nanopore native RNA sequencing. PloS One 14, e0216709 (2019).

79. Liu, H. et al. Accurate detection of m6A RNA modifications in native RNA sequences. Nature Communications 10, 4079 (2019).

80. Workman, R. E. et al. Nanopore native RNA sequencing of a human poly(A) transcriptome. Nature Methods 16, 1297–1305 (2019).

81. Parker, M. T. et al. Nanopore direct RNA sequencing maps the complexity of Arabidopsis mRNA processing and m6A modification. eLife 9, e49658 (2020).

82. Kim, D. et al. The Architecture of SARS-CoV-2 Transcriptome. Cell 181, 914–921.e10 (2020).

83. Leger, A. et al. RNA modifications detection by comparative Nanopore direct RNA sequencing. Nature Communications 12, 7198 (2021).

84. Hendra, C. et al. Detection of m6A from direct RNA sequencing using a multiple instance learning framework. Nature Methods 19, 1590–1598 (2022).

85. Wagner, J. et al. Curated variation benchmarks for challenging medically relevant autosomal genes. Nature Biotechnology 40, 672–680 (2022).

86. Aguzzoli Heberle, B. et al. Mapping medically relevant RNA isoform diversity in the aged human frontal cortex with deep long-read RNA-seq. Nature Biotechnology (2024) doi:10.1038/s41587-024-02245-9.

87. Mantel, N. & Haenszel, W. Statistical aspects of the analysis of data from retrospective studies of disease. Journal of the National Cancer Institute 22, 719–748 (1959).

88. Cochran, W. G. Some Methods for Strengthening the Common χ 2 Tests. Biometrics 10, 417 (1954).

89. Leger, A. et al. RNA modifications detection by comparative Nanopore direct RNA sequencing. Nature Communications 12, 7198 (2021).

90. Samarakoon, H. et al. Leveraging basecaller’s move table to generate a lightweight k-mer model for nanopore sequencing analysis. Bioinformatics 41, btaf111 (2025).

91. Kovaka, S. et al. Uncalled4 improves nanopore DNA and RNA modification detection via fast and accurate signal alignment. Nature Methods 22, 681–691 (2025).

92. Diensthuber, G. et al. Systematic benchmarking of dorado basecalling models for RNA modification detection with highly multiplexed nanopore sequencing. Nucleic Acids Research 54, gkag349 (2026).

93. Delgado-Tejedor, A. et al. Native RNA nanopore sequencing reveals antibiotic-induced loss of rRNA modifications in the A- and P-sites. Nature Communications 15, 10054 (2024).

94. Bosmeny, M. S. et al. A nanopore-based HIV-1 reference epitranscriptome. Nucleic Acids Research 54, gkag220 (2026).

95. Hewel, C. et al. Direct RNA sequencing enables improved transcriptome assessment and tracking of RNA modifications for medical applications. Nucleic Acids Research 53, gkaf1314 (2025).

96. Körtel, N. et al. Deep and accurate detection of m6A RNA modifications using miCLIP2 and m6Aboost machine learning. Nucleic Acids Research 49, e92–e92 (2021).

97. Arguello, A. E., Leach, R. W. & Kleiner, R. E. In Vitro Selection with a Site-Specifically Modified RNA Library Reveals the Binding Preferences of N6-Methyladenosine Reader Proteins. Biochemistry 58, 3386–3395 (2019).

98. Shi, H. et al. YTHDF3 facilitates translation and decay of N6-methyladenosine-modified RNA. Cell Research 27, 315–328 (2017).

99. Roundtree, I. A. et al. YTHDC1 mediates nuclear export of N6-methyladenosine methylated mRNAs. eLife 6, e31311 (2017).

100. Heck, A. M., Russo, J., Wilusz, J., Nishimura, E. O. & Wilusz, C. J. YTHDF2 destabilizes m6A-modified neural-specific RNAs to restrain differentiation in induced pluripotent stem cells. RNA 26, 739–755 (2020).

101. Beach, T. G. et al. Arizona Study of Aging and Neurodegenerative Disorders and Brain and Body Donation Program. Neuropathology : official journal of the Japanese Society of Neuropathology 35, 354–389 (2015).

102. nanoporetech/dorado. Oxford Nanopore Technologies https://github.com/nanoporetech/dorado (2025).

103. Li, H. Minimap2: pairwise alignment for nucleotide sequences. Bioinformatics 34, 3094–3100 (2018).

104. Li, H. et al. The Sequence Alignment/Map format and SAM-tools. Bioinformatics (Oxford, England) 25, 2078–2079 (2009).

105. De Coster, W. & Rademakers, R. NanoPack2: populationscale evaluation of long-read sequencing data. Bioinformatics 39, btad311 (2023).

106. Di Tommaso, P. et al. Nextflow enables reproducible computational workflows. Nature Biotechnology 35, 316–319 (2017).

107. Agresti, A. A Survey of Exact Inference for Contingency Tables. Statistical Science 7, 131–153 (1992).

108. Benjamini, Y. & Hochberg, Y. Controlling the False Discovery Rate: A Practical and Powerful Approach to Multiple Testing. Journal of the Royal Statistical Society. Series B (Methodological) 57, 289–300 (1995).

109. Di Tommaso, P. et al. Nextflow enables reproducible computational workflows. Nature Biotechnology 35, 316–319 (2017).

110. Kurtzer, G. M., Sochat, V. & Bauer, M. W. Singularity: Scientific containers for mobility of compute. PLOS ONE 12, e0177459 (2017).

